# The Effect of Three Key Administrative Errors on Patient Trust in Physicians: Prescription Errors, Confidentiality Breaches, and Appointment Errors

**DOI:** 10.1101/2024.12.27.24319703

**Authors:** Tetsuro Aita, Yoshia Miyawaki, Yu Katayama, Kosuke Sakurai, Nao Oguro, Takafumi Wakita, Nobuyuki Yajima, Ashwin B. Gupta, Noriaki Kurita

## Abstract

**Importance:** Understanding how administrative errors, such as prescription errors, appointment scheduling errors, and patient confidentiality breaches impact trust in physicians is crucial for improving patient-physician relationships and healthcare outcomes.

**Objectives:** To investigate the association between administrative errors, general trust in physicians and interpersonal trust in a physician.

**Design:** Cross-sectional study.

**Setting:** An online panel survey targeting adults across Japan.

**Participants:** Adults aged ≥20 years who had received treatment at least twice for non-communicable diseases within the past 6 months.

**Exposure:** Past experiences with prescription errors, confidentiality breaches, and appointment scheduling errors by personal physicians treating their non-communicable diseases.

**Primary Outcomes and Measurements:** General trust in physicians and interpersonal trust in a physician were measured using the Japanese version of the Wake Forest Physician Trust Scale.

**Results:** Among the 661 participants, nearly 14% reported experiencing at least one type of administrative error. Prescription errors were associated with a significant decrease in general trust in physicians (–9.78 points, 95% confidence interval (CI): –13.74 to –5.81). Confidentiality breaches had the most significant negative impact on interpersonal trust (–14.09 points, 95% CI: –24.35 to –3.83), followed by appointment scheduling errors (–13.56 points, 95% CI: –22.48 to –4.65). Mediation analysis revealed that the association between prescription errors and reduced general trust was partially mediated by decreased trust in personal physicians.

**Conclusions and Relevance:** Administrative errors during care for non-communicable diseases significantly undermine patients’ trust in physicians. Physicians should prioritise improving their practices, particularly regarding prescription errors, as these errors harm interpersonal relationships and have broader implications for the public’s perception of physicians.

**key messages:** *What is already known on this topic:* Patient trust in physicians has been conceptualised as two constructs: trust in personal physicians and trust in physicians in general. While administrative errors, including medication prescription errors, can cause physical harm to patients, the impact of such errors, along with issues like appointment scheduling errors and breaches of confidentiality, on trust in physicians, has not been fully explored.

*What this study adds:* This study found that prescription errors were associated with a decline in general trust in physicians. Appointment scheduling errors and breaches of confidentiality were shown to undermine interpersonal trust in personal physicians. Additionally, loss of trust in personal physicians partially mediated the association between prescription errors and loss of general trust in physicians.

*How this study might affect research, practice or policy:* Enhancing administrative practices, such as ensuring accuracy in medication prescribing, improving appointment scheduling, and safeguarding patient confidentiality, could strengthen the physician-patient relationship on a personal level and pave the way for better future interactions with other physicians.

## Introduction

Patient trust in physicians is central to medical care and plays an important role in medication adherence, adoption of recommended preventive behaviours, and ensuring the continuity of care. [1–3] Trust in physicians can be classified into two constructs: interpersonal trust, which refers to trust in a specific physician, and general trust, which pertains to trust in physicians as a collective group. [4,5] General trust is particularly important for enabling physicians to gain confidence with new patients who have limited prior interactions. [4] Although numerous studies have shown that medical errors causing physical harm result in diminished trust in the involved physician or institution, [6–8] limited understanding of how administrative errors in outpatient services—errors that do not necessarily involve physical harm—affect both interpersonal and general trust exist.

Prescription errors, for instance, can lead to the delayed recognition of adverse events, causing physical and emotional distress for patients and their families. [8,9] These errors, whether executive actions (e.g. incorrect prescriptions) or omissions (e.g. failure to prescribe), are known to impact patient well-being; however, their influence on trust in physicians remains unclear. [9] Similarly, appointment scheduling errors represent a common administrative omission, yet their impact on trust has not been extensively studied. [10] In addition, patient confidentiality breaches, a cornerstone of the doctor-patient relationship, pose another critical issue. While confidentiality breaches may not cause direct physical harm, they can erode trust, result in patient humiliation, and lead to broad disadvantages (i.e., legal sanctions). [11,12] While it is intuitive that confidentiality violations damage interpersonal trust, their potential to diminish general trust in the healthcare system has not been quantitatively examined. [12] This distinction is critical, as general trust may be impacted differently to these errors than interpersonal trust. [4,5]

Our study aimed to examine the impact of administrative errors—such as prescription errors, patient confidentiality breaches, and appointment scheduling errors—on both interpersonal trust in their physicians and general trust in physicians as a group among adults with non-communicable diseases. Furthermore, we explored whether interpersonal trust in their physicians mediates the relationship between these errors and general trust, offering insights into the importance of different dimensions of trust in healthcare management.

## Methods

### Study Design and Subjects

This cross-sectional study used an online panel survey conducted through Cross Marketing, a web-based company located in Shinjuku-ku, Tokyo, to recruit Japanese participants with non-communicable diseases. The survey was conducted on April 27 and 28, 2020, targeting individuals aged 20 years and older. Eligible participants were recruited consecutively and their responses were collected simultaneously. The study protocol was approved by the Ethics Review Board of Okayama University Hospital (No. ken-2405-041). Participants were presented with an online consent statement at the beginning of the survey, and only those who completed the survey were included in the study. This study adhered to the Strengthening the Reporting of Observational Studies in Epidemiology (STROBE) guidelines. [13]

### Screening Process and Data Collection

Quality assurance measures to mitigate random variability and enhance data reliability have been described previously. [5,14] Briefly, the survey employed multiple screening items to ensure the validity of responses. Participants were initially asked to select any non-communicable diseases for which they had received treatment at least twice within the past 6 months, with the option of selecting multiple conditions. They were then asked to identify the most burdensome diseases based on their selections. Any inconsistencies between the diseases reported in these two items led to the exclusion of the participant’s data. Furthermore, respondents were asked to list the medications prescribed for their most burdensome diseases. Any discrepancies between the most burdensome disease and appropriate medications for that disease resulted in exclusion. Two researchers independently verified the listed medications against an online database of label information and resolved any discrepancies by consensus to ensure consistency. An exception was made for respondents who selected cancer and reported “none” for prescribed medications, as the watch-and-wait approach is considered appropriate for certain cancer treatments. Additionally, a response time cut-off of 300 s, determined through prior pilot testing, was applied to exclude overly quick responses that could indicate random or non-serious answers.

### Outcome

The primary outcomes were general trust in physicians and interpersonal trust in personal physicians. Trust was measured using the Japanese version of the 5-item Wake Forest Physician Trust Scale: *Trust in Doctors Generally* and *Interpersonal Trust in Physician*. [5,15] Each item was rated on a 5-point Likert scale ranging from 1 (*totally disagree*) to 5 (*totally agree*). One negatively worded item was reverse coded, and the total score for each scale was normalised to a range of 0 to 100.

Before answering the *Interpersonal Trust in Physician* scale, respondents were instructed as follows: “Please think of the doctor who cares for your [the most troublesome disease chosen by the participants was automatically displayed here] when you answer these questions. He or she will be considered your doctor for this survey. For the next question, we are interested in your honest opinion about your doctor. Please choose the answer that best matches your opinion for each question.”

For the *Trust in Doctors Generally* scale, respondents received the following instructions: “The following questions may seem similar to the previous ones. However, they are not about your doctors, however, doctors in general. No need to be concerned if these issues have not been considered previously. No correct or incorrect responses were obtained. Please choose the answer that best matches your thoughts on doctors.”

### Exposures

Exposure included three types of administrative errors: prescription errors, appointment scheduling failures, and confidentiality breaches. Respondents were first provided with the following instructions: “Please select the response that best describes your opinion regarding the following statements about your doctor who cares for your [the most troublesome disease chosen by the participants was automatically displayed here]”. The following items were then presented: “Has your doctor ever failed to prescribe your medication, or mistakenly prescribed wrong types or dosages of your medication?”, “Has your doctor previously forgotten an outpatient or exam appointment?”, “Has your doctor ever previously discussed your illness or personal information with you in a place where someone you did not know could hear you?” For each item, respondents were asked to select either (1) (*have had*) or (2) (*have not*).

### Covariates

Potential confounders were identified based on prior literature and clinical expertise. These included age, [1,16] sex, [16] final education, household income, [16] reported disease, duration of the patient-physician relationship, [1] and patient’s general level of interpersonal traits. The patients’ general level of interpersonal trust was assessed using the 6-item General Trust Scale, [17] rated on a 5-point Likert scale ranging from *strongly disagree* to *strongly agree*. The total score was computed by summing the responses to all items.

### Statistical Analysis

Demographic and clinical variables are summarised as medians with interquartile ranges for continuous variables and frequencies with percentages for categorical variables. We used general linear models to estimate the associations between the three types of administrative errors and both interpersonal trust in a physician and general trust in physicians. Cluster-robust variance estimation was used to account for the within-prefecture clustering. [18] To assess whether trust in personal physicians mediates the relationship between specific administrative errors and general trust in physicians, we conducted a mediation analysis. A series of mediation models were fitted to examine the hypothesised relationships between administrative errors, interpersonal trust (mediator), and general trust while adjusting for the previously described covariates. The mediation analysis was performed using the user-written Stata command **“sgmediation2”**. [19] All statistical analyses were performed using Stata version 18 (StataCorp).

## Results

### Participant characteristics

A total of 964 participants met the inclusion criteria; however, 247 were excluded due to inconsistent responses regarding their most troubling disease and medication, and 46 were excluded for responding too quickly to provide logical answers. [5] Of the remaining 671 eligible participants, 661 (mean age: 63 years; 26.5% female) were included in the primary analyses and 10 were excluded due to missing data on covariates. The baseline characteristics are shown in Table 1. The most common diseases were cancer (*n* = 255; 38.6%), diabetes mellitus (*n* = 191; 28.9%), and depression (*n* = 127; 19.2%). Among the participants, 92 (13.9%) reported experiencing at least one administrative error, 72 (10.9%) experienced a prescription error, 18 (2.7%) experienced a confidentiality breach, and 26 (3.9%) reported an appointment reservation error.

**Table 1.** Participants’ characteristics.

| **Baseline Characteristics** | **Total (N = 661)** |
| --- | --- |
| **Age, years** | 63.0 (56.0–70.0) |
| **Male sex** | 486 (73.5%) |
| **Education** |  |
| Junior high school | 19 (2.9%) |
| High school | 209 (31.6%) |
| Junior college | 65 (9.8%) |
| University | 325 (49.2%) |
| Graduate school | 30 (4.5%) |
| Not answered | 13 (2.0%) |
| **Household income** |  |
| <1,000,000 yen | 40 (6.1%) |
| 1,000,000 – < 3,000,000 yen | 157 (23.8%) |
| 3,000,000 – < 5,000,000 yen | 203 (30.7%) |
| 5,000,000 – < 10,000,000 yen | 205 (31.0%) |
| ≥ 10,000,000 yen | 56 (8.5%) |
| **Patient-physician relationship duration** |  |
| <1 years | 60 (9.1%) |
| 1 – < 3 years | 212 (32.1%) |
| ≥ 3 years | 389 (58.9%) |
| **General level of interpersonal trust** | 54.2 (45.8–70.8) |
| **Reported diseases** |  |
| Arrhythmia | 37 (5.6%) |
| Ischemic heart disease | 119 (18.0%) |
| Heart failure | 15 (2.3%) |
| Diabetes mellitus | 191 (28.9%) |
| Rheumatoid arthritis/Systemic lupus erythematosus | 17 (2.6%) |
| Cancer | 255 (38.6%) |
| Depression | 127 (19.2%) |
| **The most troublesome disease** |  |
| Arrhythmia | 17 (2.6%) |
| Ischemic heart disease | 89 (13.5%) |
| Heart failure | 8 (1.2%) |
| Diabetes mellitus | 175 (26.5%) |
| Rheumatoid arthritis/Systemic lupus erythematosus | 13 (2.0%) |
| Cancer | 242 (36.6%) |
| Depression | 117 (17.7%) |

Note: Values are presented as medians (IQR) for continuous variables and n (%) for categorical variables.

### Association between trust in physicians and three types of administrative errors

After adjusting for covariates, all three types of errors were associated with decreased interpersonal trust in their physicians. As shown in Table 2 and Fig. 1, confidentiality breaches had the most significant negative impact on interpersonal trust scores (coefficient estimate: –14.09 points [95% confidence interval (CI): –24.35 to –3.83; *p* = 0.008), followed by appointment reservation errors (–13.56 points [95% CI: –22.48 to – 4.65; *p* = 0.004). Medication prescription errors also significantly reduced interpersonal trust (–5.56 points [95% CI: –10.70 to –0.41; *p* = 0.035). In contrast, general trust in physicians was primarily associated with medication prescription errors, which led to a reduction of 9.78 points [95% CI: –13.74 to –5.81; *p* < 0.001). No significant associations were found between general trust in physicians and appointment reservation errors (–6.12 points [95% CI: –12.59 to 0.35; *p* = 0.063) or confidentiality breaches (–5.23 points [95% CI: –11.33 to 0.88; *p* = 0.091).

**Fig. 1.**
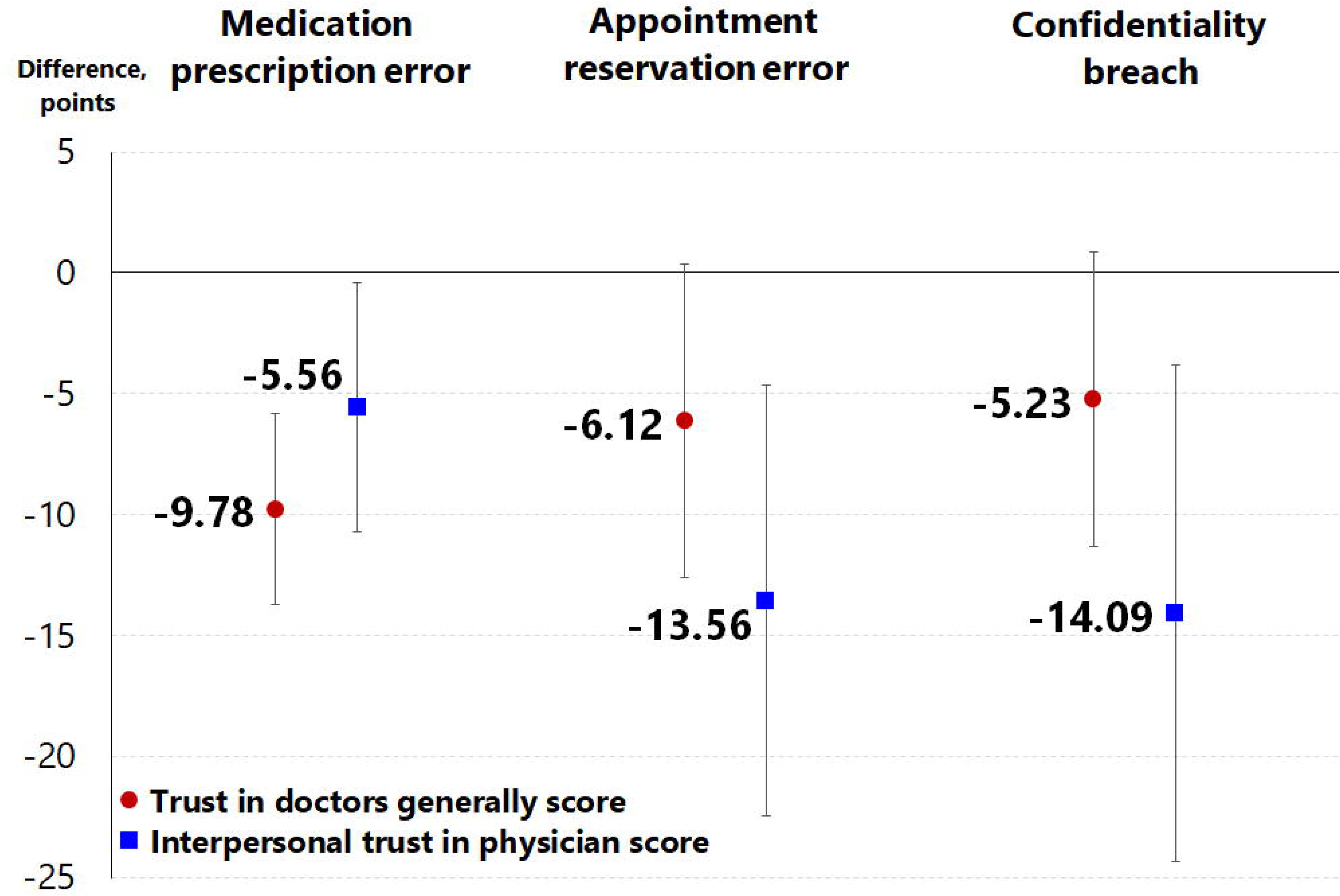
Association between Trust in physicians and administrative errors. The general linear model described in the footnotes of Table 2 was fitted to each outcome. Red circles indicate the adjusted mean difference (coefficient) in Trust in Doctors Generally scores associated with administrative errors. The blue square indicates the adjusted mean difference (coefficient) of Interpersonal Trust in Physician scores associated with administrative errors. Error bars indicate 95% confidence intervals.

**Table 2.**
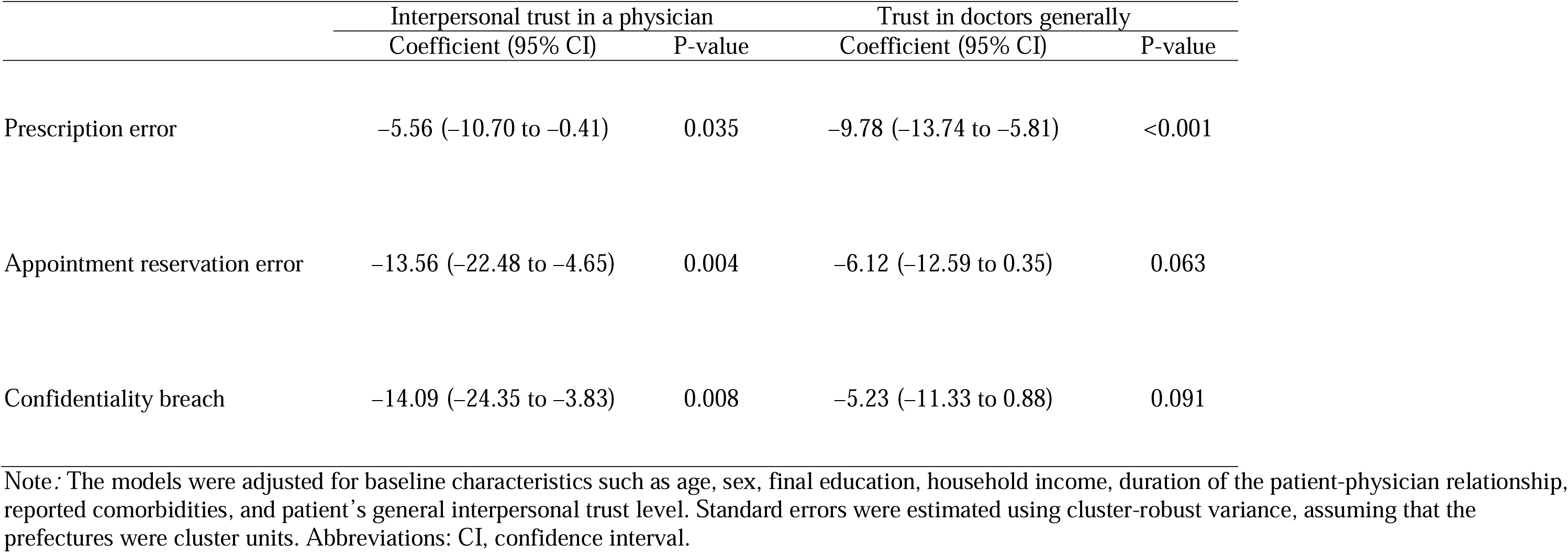
Association of administrative errors with trust in doctors generally and interpersonal trust in a physician.

### The mediating role of trust in their own physician in the association between medication prescription error and trust in general physicians

Medication prescription errors were the only type of administrative error significantly associated with decreased general trust in physicians. Both direct effects (69% of the total effect) and indirect effects (31% of the total effect) of medication prescription errors on general trust in physicians were observed. Mediation analysis revealed that a decline in interpersonal trust in their physicians partially mediated the relationship between medication prescription errors and the overall decline in general trust in physicians (Sobel test, *p* = 0.031; see Fig. 2).

**Fig. 2.**
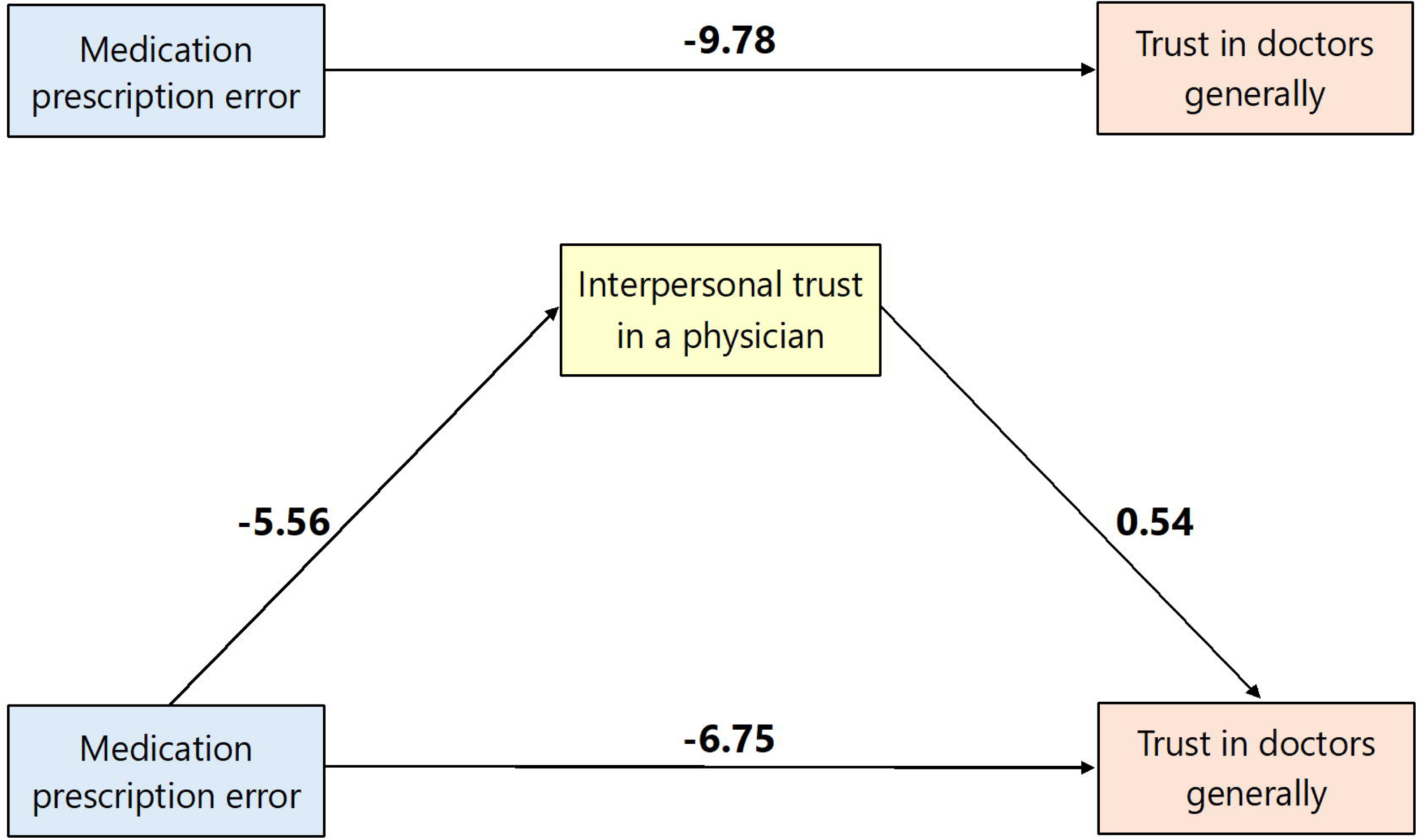
Mediating Role of Interpersonal Trust in the Association Between Medication Prescription Errors and General Trust in Doctors. This figure shows the direct and indirect paths between medication prescription errors and general trust in doctors, mediated by interpersonal trust in a physician. Direct effect: –6.75, indirect effect: –3.02 (Sobel test, *p* = 0.031). The total effect of medication errors on general trust is –9.78, with 30.9% of this effect mediated through interpersonal trust in the attending physician.

## Discussion

This study underscores the significant impact of administrative errors on patient trust in physicians among individuals with non-communicable diseases such as cancer, diabetes, and depression. Prescription errors were associated with reduced trust in both personal physicians and physicians in general, with the loss of trust in personal physicians partially mediating the loss of trust in general physicians.

Confidentiality breaches and missed appointment scheduling resulted in the greatest reduction in interpersonal trust scores, highlighting that ensuring reliable appointment scheduling is as crucial as maintaining patient confidentiality.

The reported incidence of prescription errors in primary care outpatient services affiliated with an academic medical centre in the United States is 7.6%. [20] In comparison, the prevalence of prescription errors in this study was 10.9%, focusing specifically on missed prescriptions or errors in type or dosage. One reason our percentage was slightly higher than that of the previous study is that our study relied on patient-reported reflections over a longer recall period, whereas the prior study used short-term prospective verification of prescription records. [20] Few studies have comprehensively examined the loss of trust in physicians owing to prescription errors, including their effects on general trust. Prior research quantified trust in physicians using vignettes involving adverse medication events; however, that study assessed trust based on disclosure practices rather than the presence of the error itself. [21] In this study, the reduction in trust in one’s physician due to prescription errors (5.6 points) was comparable to the impact of misdiagnosis by a different physician (4.3 points). [14] This finding suggests that prescription errors, which may be harmless, [9] are often an unexpected experience, causing negative and persistent psychological consequences. Additionally, our findings on the loss of trust in general physicians stemming from prescription errors by their personal physicians support the argument that trust in general physicians is influenced to some extent by individual experiences with their personal physicians. [4,5] In other words, prescription errors can erode trust not only in specific physicians but also in the broader healthcare system. Furthermore, the results highlighted that the loss of trust in general physicians due to prescription errors was partially mediated by diminished trust in one’s own physicians. On the other hand, the direct loss of trust in general physicians attributable to prescription errors may be shaped by the societal perceptions of physicians conveyed through the media and other social channels. [4,5,22] Particularly, exaggerated portrayals of medical errors in television dramas may contribute to negative public perceptions of healthcare systems. [22] For example, an analysis of medical errors depicted in six episodes from each of eight popular U.S. medical television series (1994–2018) revealed that medical errors occurred at a rate of 6.4 times per hour, with medication errors being the third most common. [22]

The findings indicate that, among patients with non-communicable diseases requiring ongoing management, the loss of interpersonal trust resulting from physicians discussing patient illnesses or personal information in a way that can be overheard by others, is greater than that caused by prescription errors. The issue of healthcare professionals discussing patient information in institutional spaces with limited privacy has been studied in many countries. For instance, a study conducted in five U.S. hospitals found that approximately 14% of elevator rides involved inappropriate comments from healthcare professionals. The majority of confidentiality violations, including graphic discussions of patient identifiers and treatment plans, were attributed to physicians. [23] However, breaches of confidentiality stemmed from patients’ perceptions of insufficient auditory privacy during interactions with their physicians. This issue can be attributed not only to physicians’ attitudes toward confidentiality but also to the design of consultation rooms, a problem that may be specific to Japan. Consultation rooms in Japan are often not fully walled off on all four sides. In particular, the side opposite the patient entrance is frequently left open to facilitate the movement of medical staff between consultation rooms. This design flaw allows conversations to be overheard by patients in adjacent rooms. Similar issues have been reported in a U.S. emergency room study where patients often overheard discussions about themselves and others due to insufficient space and inadequate walls on all sides. [24] Another potential source of confidentiality breaches, one not unique to Japan, may involve ward rounds during past hospitalisations, where sensitive patient information was overheard by others in neighbouring beds. [11] Unlike prescription errors, breaches of confidentiality did not significantly affect general trust in physicians in this study. This finding suggests that such breaches are viewed by patients as isolated incidents attributable to specific physicians rather than reflective of the healthcare system as a whole. [12]

The loss of trust in physicians resulting from missed appointment scheduling may be a unique issue in Japan. While follow-up appointments are commonly scheduled through electronic healthcare records in many countries, [25] this task is typically performed by the physicians themselves at the end of consultations in Japan. In contrast, in the U.S. and other countries, appointment scheduling is often handled by a hospital administrators. [10] Although clerical errors leading to missed appointments are occasionally reported globally, the administrative burden on Japanese physicians may contribute to oversights of appointment reservations. Such errors are often discovered only when patients inform their physicians. The magnitude of the loss of trust in personal physicians associated with missed appointments, comparable to that of confidentiality breaches, suggests that patients attribute these administrative errors to their personal physician rather than to systemic issues.

This study has several strengths. First, we quantified the impact of specific categories of administrative errors on trust in both one’s personal physician and general physicians, offering valuable insights into the dynamics of patient trust within healthcare settings. Second, by adjusting for patients’ general tendency to trust others as a covariate, the analysis isolated the impact of these errors on trust in physicians independently of general trustworthiness.

However, this study has certain limitations, which must be acknowledged. First, the classification of administrative errors relied on participants’ self-reports, which may have introduced misclassification. For instance, confidentiality breaches may have been underestimated due to the reliance on participants’ recollections of past interactions. Second, the study did not examine the specifics of medical errors, such as the type of medication error (e.g. dosage exceeding label instructions or misprescription of a medication with a different indication) or the consequences of these errors (e.g. the presence, or absence, or severity of adverse events).

In conclusion, this study provides unique insights into how administrative errors, such as prescription errors, missed appointment scheduling, and confidentiality breaches, directly and negatively impact the quality of physician-patient interactions. Addressing these issues is crucial for fostering strong physician-patient relationships and improving outcomes, particularly in the long-term management of Japanese adults with non-communicable diseases.

## Data Availability

The data supporting the findings of this study are available from the corresponding author upon request.

## Ethical Approval

The study protocol was approved by the Ethics Review Board of Okayama University Hospital (No. ken-2405-041).

## Author Contributions

Aita, Miyawaki, and Kurita had full access to all data in the study and took responsibility for the integrity of the data and the accuracy of the data analysis.

*Concept and design*: Aita, Kurita.

*Acquisition, analysis, or interpretation of data*: All authors.

*Drafting of the manuscript*: Aita, Miyawaki, Kurita.

*Critical revision of the manuscript for important intellectual content*: All authors.

*Statistical analysis*: Aita, Miyawaki, Kurita.

*Obtained funding*: Kurita

*Administrative, technical, or material support*: Wakita, Yajima

*Supervision*: Yajima, Wakita, Gupta

## Additional Contributions

TA, YK, YM, TF, NY, AG, and NK designed and conceived the study. RS, KS, and NO collected data. TA, YK, YM, AG, and NK analysed and interpreted the results and drafted the manuscript. All the authors have read and approved the final version of the manuscript.

